# Social Validation of Post-Treatment Outcomes of Adults Who Stutter Who Participated in the Blank Center CARE^™^ Model Treatment: Replication and Extension

**DOI:** 10.1101/2024.10.19.24315802

**Authors:** Geoffrey A. Coalson, Courtney T. Byrd

**Affiliations:** The University of Texas at Austin

**Keywords:** stuttering, adults, treatment, social validation, non-ableist

## Abstract

**Purpose:** Social validation of treatment outcomes provides external validity of treatment from the perspective of untrained observers. To date, clinical efficacy studies of the Blank Center CARE Model indicate post-treatment gains in communication competence from the perspective of the participants and clinicians. An initial social validation study corroborated positive self- and clinician-rated treatment ratings with ratings from the general public for a single participant. The present study was designed to replicate and extend these findings by assessing clinical outcomes from the perspective of untrained observers across multiple participants and contexts.

**Method:** Ten adults who stutter provided communication samples one week before and after completing the Blank Center CARE Model treatment. A total of 1,110 untrained observers were recruited. Each untrained observer rated only one participant at one timepoint (pre-treatment or post-treatment) in one context (dyad or presentation), and each participant was asked to provide only one rating (communication competence or stuttering severity).

**Results:** A significant interaction indicated that post-treatment gains were observed for communication competence, but not stuttering severity, for both contexts.

**Conclusions:** Present findings provide further social validation of the Blank Center CARE Model of treatment. Untrained observers confirmed that participation in this strengths-based approach significantly enhances communication competence. Notably, these changes were observed regardless of pre- to post-treatment stuttering severity, lending additional support to the premise that communication and fluency are independent constructs.

## Social Validation of Post-Treatment Outcomes of Adults Who Stutter Who Participated in the Blank Center CARE Model Treatment: Replication and Extension

A core component of the Blank Center CARE^™^ Model for stuttering (Communication, Advocacy, Resiliency, Education, hereinafter the CARE Model) is improving the communication competence of individuals who stutter while excluding any clinical goals designed to change stuttering frequency or severity. Training in communication competence for individuals who stutter, as conceived within the CARE Model, includes explicit training in core communication skills and, importantly, considers communication to be effective only when participants stutter openly. Positive post-treatment gains in communication competence for adults who stutter have been reported from the perspective of clinicians (Byrd et al., 2022) and the participants themselves (Coalson et al., 2024). Social validation studies provide external confirmation of treatment outcomes observed by individuals in the general public with no vested interest or knowledge of the expected treatment outcomes (Kazdin, 1999; Schloss et al., 1987).

A recent social validation study by Byrd et al. (2024a) asked untrained observers to rate the communication competence of an adult who stutters based on one video sample recorded either pre-treatment or post-treatment. The preliminary findings found that significant clinician-rated and self-rated gains in communication competence reported in previous studies (Byrd et al., 2022; Coalson et al., 2024) were also observed from the perspective of the general public.

As noted in Byrd et al. (2024a), significant post-treatment gains were limited to a single participant in a single speaking context (i.e., dyadic mock interviews). Further, although stuttering severity ratings of the selected pre- and post-treatment videos were carefully controlled and comparable, comparison of untrained observers’ ratings of communication competence versus ratings of stuttering severity were not included during analysis. The purpose of this study, therefore, was to replicate and extend findings in Byrd et al. (2024a) by (a) increasing the number of participants (*N* = 10, as opposed to *N* = 1) and raters (*N* = 1,110, as opposed to *N* = 81), (b) including ratings of communication competence and stuttering severity (as opposed to communication competence alone), and (c) rating video samples from two different speaking contexts - dyad and presentation (as opposed to dyad alone).

## Stuttering treatment for adults

Clinical studies examining outcomes for adults who stutter have traditionally focused on fluency-focused treatments (see systematic reviews by Baxter et al., 2016; Johnson et al., 2016). Across these studies, post-treatment gains in fluency have been observed, but tempered by individual variability, relapse, and participant dissatisfaction (e.g., Arya & Geetha, 2013; Craig & Hancock, 1995; Cream et al., 2003; Irani et al., 2012; National Stuttering Association, 2009; Stewart & Richardson, 2004; Yaruss et al., 2002). Additional research has not found that treatments that target fluency yield improved communication competence (speaker-perspective: Constantino et al., 2020; Corcocan & Stewart, 1995; Cream et al., 2003; Stewart & Richardson, 2004; listener-perspective: De Nardo et al., 2023; Manning et al., 1999; Von Tiling, 2011) or self-reported quality of life (e.g., Boyle, 2015; Byrd, 2021). Notably, the present authors have reported (e.g., Byrd et al., 2021; Byrd et al., 2022; Coalson et al., 2024; Byrd et al., 2024b) – based on the perspective of people who stutter, clinicians and untrained observers – that changes in fluency are not required for significant changes in the communication effectiveness and/or quality of life of children and adults who stutter. These preliminary findings provide support for the CARE Model of treatment, an alternative approach to stuttering treatment that is consistent with the recent focus on mitigating ableism (i.e., fixing or curing a disabling condition) in national health care policies held by the National Institutes of Health National Advisory Board on Medical Rehabilitation Research (NIH-NABMRR, 2022) and the NIH Advisory Committee Subgroup on Individuals with Disabilities (NIH, 2022), as well as the revised policies of the American Psychological Association that move away from ableist practices during clinical design and practice (APA, 2022).

Specifically, the CARE Model (Byrd, 2023b; Byrd et al., 2024c) provides a manualized, strengths-based approach to stuttering treatment that targets communication competence, advocacy, resiliency, and education – and that does not aim to fix or cure stuttering through the targeting of goals designed to directly or indirectly increase fluency and/or modify stuttered speech (for summary of CARE Model treatment, see Appendix A). Clinical studies (children: Byrd et al., 2021, Byrd et al., 2024b; adults: Byrd et al., 2022, Coalson et al., 2024) indicate significant post-treatment improvements in communication competence that are independent from stuttering severity.

## Clinician ratings

For example, Byrd et al. (2021) examined 37 children who stutter before and after participation in CARE Model treatment. A certified and licensed speech-language pathologist (SLP) who was not trained in this approach and was not knowledgeable of the targets or intended outcomes, rated nine specific communication competences (i.e., language use, language organization, speech rate, intonation, volume, gestures, body position, eye contact, and facial affect) during impromptu presentations provided pre- and post-treatment. In addition to replicating the cognitive and affective gains in previous studies completed with children who stutter (Byrd et al., 2016a, 2016b, 2018), significant post-treatment gains were observed for eight of these nine competencies. The gains in communication competence observed by clinicians were replicated in a larger cohort (Byrd et al., 2024b; *N* = 61), with significant gains observed for seven of these nine communication competencies. Importantly, post-treatment gains were not significantly associated with pre-treatment stuttering frequency (Byrd et al., 2021; Byrd et al. 2024b). Similar to findings with children, Byrd et al. (2022; *N* = 11) found that an unfamiliar clinician rated adults who stutter significantly higher in eight of the nine communication competences following CARE Model treatment, and these changes were observed by the clinician independent of stuttering severity, suggesting that pre-treatment stuttering severity does not predict post-treatment communication competence. Taken together, these findings suggest that CARE Model treatment is effective in increasing perceived communication competence irrespective of stuttering severity, as rated by clinicians who are not trained in expected outcomes of this approach.

## Self ratings

Clinician-rated gains in communication competence following CARE Model treatment have also been reported from the perspective of the participant. For example, Byrd et al. (2022; *N* = 9) examined self-rated communication competence of adults who stutter following CARE Model treatment using the Self-Perceived Communication Competence scale (SPCC; McCrosky & McCroskey, 1988), which measures participants’ self-rated communication abilities across seven different speaking scenarios (e.g., dyad, small group, large presentation). Post-treatment SPCC scores improved but did not reach significance (*p* = .31) in this small sample. A larger follow-up study by Coalson et al. (2024; *N* = 33), however, found significant post-treatment gains in communication competence, as measured by the SPCC. As was the case with the clinician ratings, these gains were not accompanied by significant changes in stuttering severity post-treatment.

Collectively, both clinician- and self-rating of communication competence following CARE Model of treatment indicate improvements in communication competence are not dependent on changes in stuttering behaviors. Although these self- and clinician-rated outcomes are critical to our understanding of the clinical efficacy of this treatment program, ratings of unfamiliar observers, or rather the general public, are needed to provide corroborating evidence that the reported gains are not limited to the perspective of those involved in the treatment.

## Unfamiliar observer ratings

Social validation studies provide a third-party evaluation of treatment measures from the perspective of unfamiliar individuals who are not invested nor aware of the intended clinical outcomes (Kazdin, 1977; Finn & Sladeczek, 2001). For example, Schloss et al. (1987a) conducted a study to assess whether assertiveness training provided to three adults who stutter resulted in changes that were observable not only to the clinicians, but to a small cohort of untrained observers (*N* = 10 graduate students). Each student randomly viewed only one pre- or post-treatment video sample of an adult who stutters and rated their overall assertiveness during a mock interview. Because findings indicated an increase in assertiveness in videos recorded after treatment, relative to pre-treatment videos, the researchers extended findings observed within the clinic to the general public. A second social validation study conducted by Schloss et al. (1987b) replicated these findings, again instructing untrained observers (*N* = 11) to rate video samples of three adults who stutter pre- and post-treatment in a dyadic interview. Notably, post-treatment gains in assertiveness ratings were not consistently associated with stuttering severity across these studies.

Werle and Byrd (2022a, 2022b) conducted a experimental study wherein untrained observers, in this case university professors (*N* = 158, 238), rated the communication competence of an adult actor simulating stuttering and also demonstrating high or low communication competence behaviors (e.g., facial affect, vocal variety, gestures, body positioning). Each untrained observer rated only one video, and each rated the presenter using a standardized rubric of ‘delivery’ (i.e., communication behaviors including vocal variety, eye contact, engagement, gestures). Although stuttering was present and controlled across videos (i.e., 15% frequency, identical type and secondary behaviors), professors rated the presenter with high communication competence significantly higher than low communication competence. Although the presenter was an actor simulating stuttering, rather than an individual who stutters who had undergone treatment, findings provide some external validation of the importance of communication competence to untrained observers relative the stuttering frequency or severity.

Byrd et al. (2024a) conducted a social validation study to assess whether untrained observers corroborated the clinician-rated and self-rated gains in communication competence of an actual adult who stutters following CARE Model treatment. In this study, 81 untrained observers viewed one video - either a pre-treatment or post-treatment video of an adult who stutters during a one-on-one mock interview. After viewing a video, untrained observers were asked to rate the communication competence of the interviewee. Findings indicated that untrained observers rated the adult who stutters as a significantly stronger communicator in post-treatment videos. Notably, stuttering of the participant in video stimuli were rated by clinicians as similar in terms of frequency (pre-treatment – 7%, post-treatment – 6%, as rated by Yairi and Ambrose [2005] guidelines) and severity (pre-treatment – Moderate [score = 27], post-treatment [score = 25], as rated by the Stuttering Severity Instrument-Fourth Edition, Riley [2009]).

Stuttering severity was also rated as similar by a separate group of untrained observers using a 100-point visual analog scale (*N* = 120, pre-treatment: *M* = 64.71; post-treatment: *M* = 64.95; *p* =.95). Although post-treatment differences in stuttering were nominal from the perspective of clinicians and untrained observers, a supplemental replication analyses was nevertheless completed within the same study (Appendix S3, *N* = 96 untrained observers) based on pre- and post-treatment videos from a second adult who stutters with an inverse stuttering profile – that is, increased stuttering post-treatment - as rated by clinicians (pre-treatment stuttering frequency and severity: 5%, Mild [Total SSI-4 score = 10]; post-treatment frequency and severity: 9%, Mild [Total SSI-4 score = 14]) and untrained observers (*N* = 85; pre-treatment: 27.19; post-treatment: 29.07, *p* = .65). Similar to the primary analyses, untrained observers rated communication competence in the post-treatment video sample to be significantly stronger than the pre-treatment video sample. Further, post-treatment gains in communication competence in the primary and supplementary analysis remained significant when observer-based factors (e.g., demographics, familiarity with a person who stutters) were statistically controlled.

## Purpose of the present study

The design of Byrd et al. (2024a), wherein many untrained observers viewed and rated video samples from a single adult who stutters pre- and post-treatment, allowed researchers to capture the variance of responses amongst the general public. This provided social validation of one of the primary clinical outcomes of the CARE Model – increased communication competence with no attempts to modify stuttered speech - for one participant. Byrd et al. (2024a) noted that the gains reported from the perspective of untrained observers viewing only one participant may or may not be observed when evaluating a group of participants before and after treatment. Finally, Byrd et al. (2024a) also noted that by restricting the context of the video sample to dyadic interaction (i.e., mock interview format), the communication gains could not be generalized to additional challenging speaking contexts for adults who stutter. Replication of these findings for one participant in a larger cohort of adults who stutter would provide evidence that treatment effects, as rated by the untrained observer, generalize across a variety of participants. In addition, although stuttering severity was held constant in pre- and post-treatment video samples in Byrd et al. (2024a), and both supplemental replication data (in Byrd et al., 2024a, Appendix S3) and qualitative data (in Byrd et al., 2024a, Appendix S2) supported the notion that communication competence and stuttering severity were described as separate constructs by untrained observers, one might argue that ratings of communication competence, nevertheless, will reflect ratings of stuttering severity in a larger cohort. The purpose of the present study was to directly address these three factors by replicating and extending the design of Byrd et al. (2024a).

First, to capture the variance of treatment effects across participants, untrained observers in the present study rated a larger number of adults who stutter before and after treatment (*Timepoint*). If the perception of communication competence by untrained observers from a larger cohort of clients replicates post-treatment gains observed for one client in Byrd et al. (2024a), the findings will provide broader social validation of the intended treatment effects across multiple participants. Second, to examine whether ratings of communication competence and stuttering severity of a larger participant cohort were perceived as separate constructs by the general public, the video samples were rated for stuttering severity as well as communication competence (*Condition*). If untrained observers provide dissimilar ratings for stuttering and communication across multiple participants, such findings would suggest that stuttering and communication are considered distinct constructs by the general public. Third, to capture the variance of treatment effects across contexts, untrained observers viewed pre- and post-treatment videos of each participant in two different speaking situations: dyadic interaction and oral presentations (*Context*). If untrained observers report that changes in post-treatment communication competence extend across contexts, such outcomes will indicate the treatment effects of the CARE Model can be generalized beyond a single interaction style.

In sum, the present study will serve as a large-scale social validation study of the CARE Model treatment program described in Byrd et al. (2024a), wherein changes in communication competence are targeted in the absence of fluency goals, but rather goals designed to guide adults to communicate effectively while stuttering openly. Specifically, we are examining the following research question: Does communication competence and stuttering severity differ pre- versus post-treatment from the perspective of untrained observers?

## Method

The following study was approved by the Institutional Review Board at the University of Texas at Austin (IRB: 2015-05-0044; see also Byrd, 2023a). All participants depicted in the video stimuli provided written consent prior to enrollment in treatment. None of the participants had previously received CARE Model treatment. Table 1 provides demographic and treatment history details for each participant.

**Table 1:**
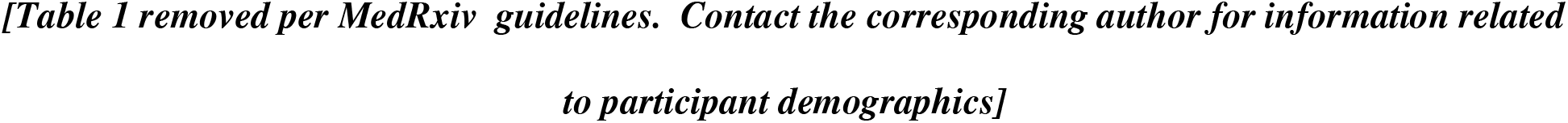
Demographics of Participants

Forty video samples (10 participants x 2 contexts x 2 timepoints) were embedded in a Qualtrics-based survey and distributed to untrained observers via the Prolific^©^ online data collection platform. Untrained observers were recruited from over 83,000 adults in the United States, United Kingdom, Australia, and Canada. All participants were paid for their participation in accordance with standards of distribution for Prolific^©^ ($1.80 per participant, *Mdn* = 9 min per survey; calculated on $12/hour base rate). Participants who elected to participate in the study were first presented with an informed consent cover letter that described the purpose of the study, compensation procedures, the right to discontinue participation, and IRB approval information. Consent to participate was acknowledged by clicking the advance button.

Each participant was randomly assigned to one of 40 videos (10 participants x 2 timepoints x 2 context). Participants were excluded from participation if they had completed a survey associated with the other 39 video samples. Each survey including a dyadic interaction began with the following instructions: *You are about to watch a video of an interview. Immediately following the video, you will be asked questions about the interviewee. The interview will be approximately 5 to 7 minutes in length. You will only be able to move forward in the survey after you have watched the video in its entirety*. Each survey including an oral presentation began with the following instructions: *You are about to watch a video of a presentation. Immediately following the video, you will be asked questions about the presenter. The presenter was instructed to give an impromptu Zoom-based presentation to a small audience on one of five topics (i.e., new inventions, best advice, best purchase, favorite month, favorite historical figure). The presenter was given 1 minute to prepare. The interview will be approximately 2 to 3 minutes in length. You will only be able to move forward in the survey after you have watched the video in its entirety*. Following the video, respondents were provided only one of the two following instructions accompanied by a 0-100 visual analog rating scale (VAS): (1) *Using the scale below, please rate the communication competence of the person. 0 = Communication skills not at all effective, 100 = Communication skills extremely effective.*(2) *Using the scale below, please rate the stuttering severity of the person. 0 = no stuttering, 100 = extremely severe stuttering*.

As in Byrd et al. (2024a), participants were then asked to provide demographic information (e.g., age, race, ethnicity, gender, education, occupation, primary language). Participants also provided information about their personal relationship with stuttering, persons who stutter, or other communication disorders (*Do you personally know a person who stutters? If so, please describe your relationship and how long you have known this person. Have you had previous speech, language, and/or hearing evaluation or therapy?*) as well as any visible or nonvisible diagnoses unrelated to communication difficulties. Appendix B provides a detailed demographic summary of untrained observers.

Consistent with Prolific^©^ policy, and to improve response fidelity (Peer et al., 2021), each survey included two comprehension checks and two attention checks (Oppenheimer et al., 2009). Multiple-choice comprehension check screening items were included to ensure that participants understand critical elements of the survey (e.g., understanding of compensation terms, understanding of instructions). Multiple-choice attention check items included instructional manipulation checks (e.g., *When asked to select your favorite color, select green*.

*This is an attention check. What is your favorite color?*) and nonsensical questions (e.g., Rate your agreement with the following statement. *I swim across the Atlantic Ocean to get to work everyday*). Participants must respond to both comprehension check and attention check questions correctly to proceed. The survey was initiated by 1,141 respondents. Of these 1,141 respondents, 31 (2.72%) did not pass the attention or comprehension check questions during the survey and were excluded from final analysis. The final corpus included 1,110 untrained observers as respondents (see Appendix B for demographic summary of untrained observers).

## Procedures

Three primary variables were examined in the present study: Timepoint, Context, and Condition. *Timepoint* was defined as whether the video sample was recorded: (1) pre-treatment (i.e., one week prior to the first treatment session) or (2) post-treatment (i.e., one week following the final treatment session). *Context* was defined as the sampling context of the video: (1) a dyadic interaction (i.e., virtual mock interview with an unfamiliar interviewer), or (2) an oral presentation (i.e., virtual impromptu oral presentation on one of five topics to two additional audience members). *Condition* was defined as which perceptual rating the untrained observers were asked to provide: (1) a rating of communication competence, or (2) a rating of stuttering severity. After viewing one video, each participant was asked to rate either (a) communication competence, or (b) stuttering severity, using a 100-point VAS described above. Similar to Byrd et al. (2024a), each untrained observer viewed and rated only one video, thereby eliminating order effects, and were blinded to the time point of the video they observed. Table 2 provides a summary of the number of participants who rated each video stimuli used in the present study.

**Table 2:** Number of Non-Overlapping Untrained Observers Serving as Raters for Each Video Sample (Condition x Timepoint x Context) Across Participants

| Communication Competence |  | Stuttering Severity |  |
| --- | --- | --- | --- |
| Pre-Treatment | Post-Treatment | Pre-Treatment | Post-Treatment |

| P | Dyad | Pres | Dyad | Pres | Dyad | Pres | Dyad | Pres | *N* |
| --- | --- | --- | --- | --- | --- | --- | --- | --- | --- |
| 1 | 17 | 12 | 12 | 12 | 15 | 12 | 12 | 14 | 106 |
| 2 | 16 | 12 | 15 | 15 | 13 | 15 | 13 | 13 | 112 |
| 3 | 15 | 15 | 13 | 13 | 15 | 13 | 11 | 13 | 108 |
| 4 | 10 | 13 | 16 | 15 | 14 | 14 | 13 | 15 | 110 |
| 5 | 13 | 12 | 14 | 12 | 13 | 13 | 15 | 14 | 106 |
| 6 | 14 | 16 | 14 | 15 | 12 | 16 | 15 | 14 | 116 |
| 7 | 14 | 16 | 15 | 16 | 12 | 11 | 15 | 14 | 113 |
| 8 | 15 | 16 | 12 | 14 | 14 | 12 | 14 | 15 | 112 |
| 9 | 15 | 16 | 13 | 14 | 15 | 14 | 15 | 15 | 117 |
| 10 | 12 | 16 | 15 | 14 | 12 | 13 | 14 | 14 | 110 |
| *N* | 141 | 144 | 139 | 140 | 135 | 133 | 137 | 141 | 1,110 |

*Note.* P = Participant; Pres = presentation.

## Analysis

A three-factor ANOVA was conducted to compare untrained observers’ ratings, with Timepoint (Pre-Treatment, Post-Treatment), Condition (Communication Competence, Stuttering Severity) and Context (Dyad, Presentation), as the three independent categorical factors and rating on the 100-point VAS as the continuous dependent variable. The three-way ANOVA was two-tailed (α = .05) and Bonferroni-adjusted p-values were applied during planned comparisons. Effect sizes were calculated and interpreted using ηρ^2^ (Cohen, 1988).

To account for the potential influence of listener-based variables, similar to Byrd et al. (2024a), nine additional observer-based factors were included as covariates: (1) age, (2) race identification, (3) ethnicity identification, (4) gender identification, (5) years of education, (6) primary spoken language, (7) knowing a person who stutters, (8) number of years the observer has known a person who stutters, and (9) visible and/or mixed disability. Race and gender categories had an insufficient number of respondents for valid statistical analysis across groups. Rather than exclude demographic categories with a limited number of respondents, and similar to Byrd et al. (2024a), groups were either combined or redistributed. Race categories with ≤10 (Native American or Alaskan Native, Native Hawaiian or Other Pacific Islander, Race Not Described) were re-grouped into a single category. Survey respondents who identified as non-binary were randomly assigned and evenly redistributed to male or female.

## Results

A significant main effect was detected for Timepoint *F*(1,1109) = 5.04, *p* = .025, ηρ2 = .005 [small effect size], Condition *F*(1,1109) = 92.66, *p* < .001, ηρ2 = .078 [large effect size], and Context *F*(1,1109) = 5.08, *p* = .024, ηρ2 = .004 [small effect size]. No significant two-way interaction was detected for Timepoint x Context *F*(1,1109) = 2.78, *p* = .097 nor Condition x Context *F*(1,1109) = .09, *p* = .765. A significant two-way interaction was detected between Timepoint and Condition *F*(1, 1109) = 7.19, *p* = .007, ηρ2 = .007 [medium effect size]. As depicted in Figure 1, significant pre- to post-treatment gains were observed for communication competence (*p* < .001; dyad: *p* = .004; presentation: *p* = .035) but not stuttering severity (*p* = .757; dyad: *p* = .298; presentation: *p* = .137). Findings also indicated that, irrespective of context, significant differences were observed between ratings of communication competence and stuttering severity at pre-treatment (*p* < .001; dyad: *p* < .001; presentation: *p* < .001) and post-treatment (*p* < .001; dyad: *p* < .001; presentation: *p* < .001), suggesting that untrained observers perceived these to be distinct variables. No significant three-way interaction was detected *F*(1, 1108) = .71, *p* = .401.

**Figure 1:**
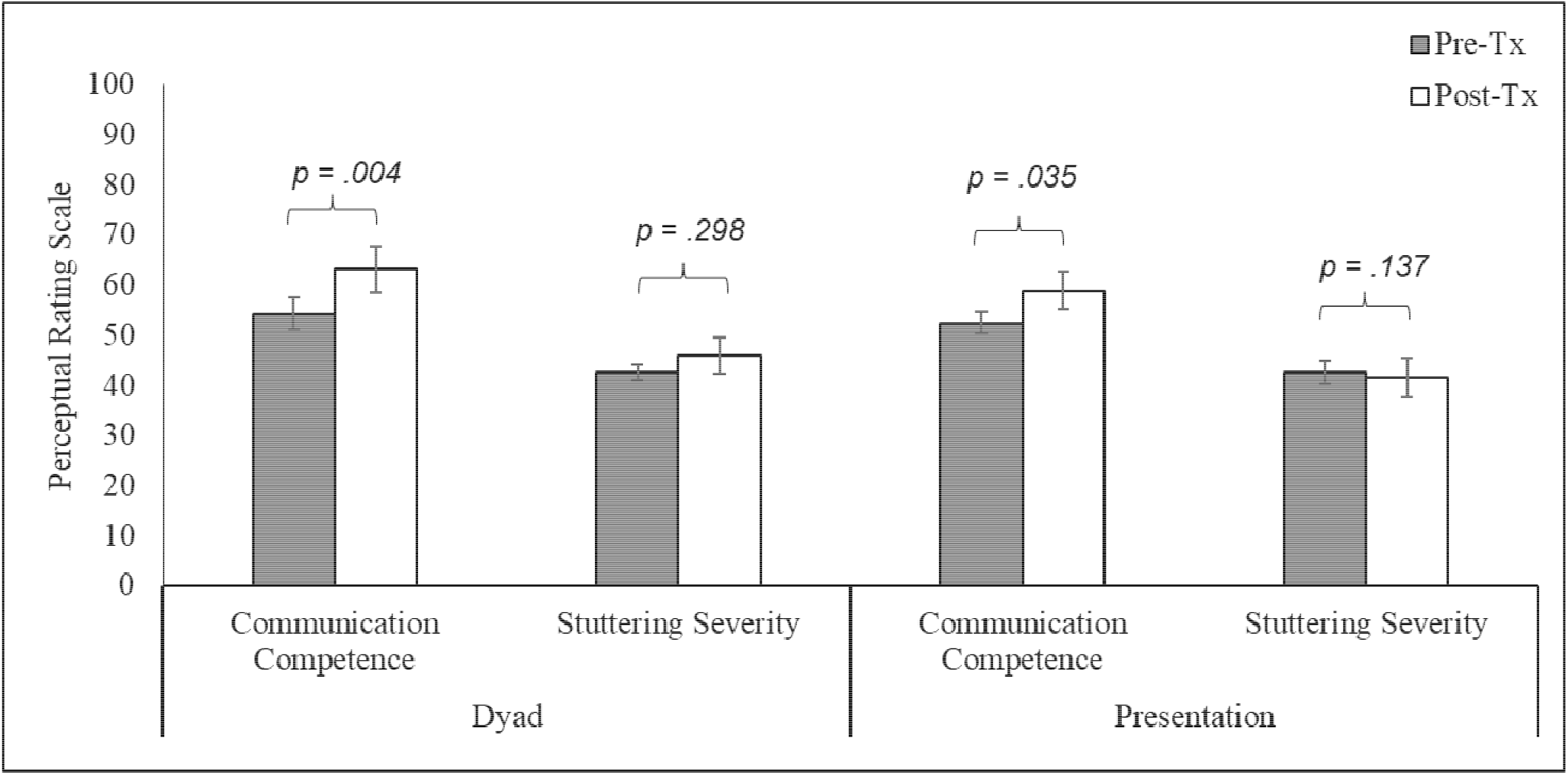
Ratings of Communication Competence and Stuttering Severity for Adults Who Stutter at Before and After Treatment (Pre-Tx, Post-Tx) Across Speaking Contexts (Dyad, Presentation)

Of the nine observer-based factors included as covariates, three were identified as significant: age *F*(1, 1109) = 6.30, *p* = .012, ηρ2 = .006 [medium effect size]; ethnicity *F*(1, 1109) = 13.45, *p* < .001, ηρ2 = .012 [large effect size]; personally knowing an adult who stutters *F*(1, 1109) = 5.68, *p* = .017, ηρ2 = .005 [small effect size].

## Discussion

The purpose of this social validation study was to assess whether gains observed following CARE Model treatment by clinicians and participants were also observed by untrained observers. This study replicates and extends a previous social validation study (Byrd et al., 2024a) in terms of number of participants (*N* = 10, as opposed to *N* = 1), raters (*N* = 1,110, as opposed to *N* = 81), context (dyadic exchanges and oral presentations, as opposed to dyad alone), and perceptual ratings provided by untrained observers (communication competence and stuttering severity, as opposed to communication competence alone). Findings indicate significant gains in communication competence pre- to post-treatment across contexts with no significant changes in stuttering severity.

## Communication Competence

Untrained observers rated the communication competence of adults who stutter as significantly higher for videos recorded after CARE Model treatment compared to videos recorded before treatment. These findings replicate ratings from untrained observers in Byrd et al. (2024a, *N* = 81 raters, *N* = 1 adult who stutters) on a larger scale (*N* = 1,110 raters, *N* = 10 adults who stutter). Significant gains post-treatment also corroborated post-treatment gains in previous studies observed by participants (Coalson et al., 2024, *N* = 33) and clinicians (Byrd et al., 2022, *N* = 11). Together, outcomes suggest that the communication competence of adults who stutter following CARE Model treatment are perceived as significantly improved from multiple perspectives.

As noted, social validation can provide evidence that self-perceived or clinician-perceived gains following treatment are not based on potentially biased perspectives. It is important to note that although the general public in the present study provide corroborating evidence of post-treatment gains in communication competence, the perspective of the general public should never take priority over participant self-perception during clinical intervention. That is, these data should be considered supporting evidence for the treatment, but not used as a criterion for clinical success. Failure to make this distinction risks a scenario wherein the general public, or perhaps clinicians, promote treatment outcomes they consider to be successful even when the participant does not (e.g., Constantino et al., 2017; Cream et al., 2003; Stewart & Corcoran, 1995).

## Stuttering Severity

The present study also asked untrained observers to rate videos based on stuttering severity. As noted, no significant changes in untrained observers’ ratings of stuttering severity. This finding replicates Byrd et al. (2024a) who reported significant changes in communication competence of videos that did not statistically differ in stuttering severity, as rated untrained observer (primary analysis, *N* = 81 raters, *p* = .95; supplemental replication analysis, *N* = 96 raters; *p* = .65). Findings also replicate data from observers in Werle and Byrd (2022a, 2022b) and self-ratings in Coalson et al. (2024), in that gains in communication competence for adults who stutter can be observed with no changes in stuttering severity.

Unlike the present study, videos in Byrd et al. (2024a) were selected specifically to control for pre- and post-treatment variance in stuttering severity when assessing untrained observers’ ratings of communication competence. In the present study, pre- and post-treatment stuttering severity were allowed to freely vary across participants. Inclusion of these ratings during analyses provided the opportunity to contrast observer ratings of stuttering severity with communication competence. At both timepoints and irrespective of context, untrained observers provided significantly different ratings for communication competence and stuttering severity, suggesting that these were considered to be independent constructs. Together, findings provide large-scale social validation for a core clinical goal of the CARE Model - improved communication competence without changes in stuttering severity.

## Speaking Context

An extension of Byrd et al. (2024a) was the assessment of speakers in two different speaking contexts - dyadic interaction (i.e., mock interview) and oral presentation (i.e., impromptu presentation to a small group). Untrained observers rated participants similarly - that is, significant gains in communication competence with no significant changes in stuttering severity - in both contexts. Findings from dyadic interactions replicate findings based on mock interviews in Byrd et al. (2024a), and extend these gains to a second, dissimilar speaking context (i.e., oral presentation). Findings suggest that changes in communication are not restricted to the communication demands of one-on-one dyadic interactions and can be generalized across contexts.

Findings inform clinical outcomes reported by adults who stutter, as measured by the SPCC, in Coalson et al. (2024). As noted, the SPCC provides an index of self-perceived communication competence across a variety of speaking contexts (i.e., public presentation, large meeting, group interaction, dyadic interaction) and audience types (i.e, stranger, acquaintance, friend). Although a priori power analysis in Coalson et al. (2024) restricted pre- to post-treatment comparison to the Total SPCC score (i.e., averaged across seven context-dependent subscales), descriptive data indicated post-treatment gains communication competence for speaking contexts similar to those rated by untrained observers in the present study. Specifically, a greater percentage of adults rated themselves as having “high” communication competence (> 87 Total SPCC Score) during dyadic interactions (pre-treatment: 21%, post-treatment: 36%) and presentations (pre-treatment: 30%, post-treatment: 48%) and with strangers (pre-treatment: 24%; post-treatment: 61%). Significant post-treatment gains found in the present study from the perspective of untrained observers, along with the descriptive gains from the perspective of the participant in Coalson et al. (2024), suggest that treatment effects generalize across contexts.

## Observer-Based Factors

Three observer-based factors were found to influence perceptual ratings: age, ethnicity, and whether the untrained observer personally knows a person who stutters. In general, younger participants and participants who self-identified as non-Hispanic or Latino provided higher ratings of communication competence than older and/or Hispanic or Latino observers. These ratings, however, were not specific to either time point or either speaking context. Comparison across studies provide some insight into the relative consistency of the factors when evaluating individuals who stutter. For example, the two demographic factors - age and ethnicity - were not identified as influential factors on ratings of communication competence in Byrd et al. (2024a) during primary analysis, or supplemental replication analysis. Previous research has identified age as a significant factor in the evaluation of negative beliefs or attitudes towards stuttering (e.g., Arnold et al., 2016; Valente et al., 2017). Recent studies have also explored the beliefs of Hispanic/Latino communities towards stuttering (e.g., Dean & Medina, 2021; Medina et al., 2024). A recent study by Young and Byrd (2024), however, found that age and ethnicity did not significantly influence employers’ beliefs about individuals who stutter or their perception of the communication competence after watching video samples of a job candidate who stutters. The exact reasons for these changes are speculative at this point, and the purpose of including these factors as covariates was to control for observer-based factors of significance, should they be observed. It should be noted that findings across studies may be inconsistent due to the overall lack of cultural diversity in stuttering research. Nevertheless, as the range of potentially influential demographic factors continues to grow, researchers should continue to collect detailed demographic data in studies of clinical outcomes. Importantly, although a variety of demographic variables may influence ratings in studies of treatment efficacy from the perspective of the general public, the positive gains of treatment remained in the present study significant after controlling for these factors.

Familiarity with an adult who stutters, however, has been a reliably influential factor when examining perceptual ratings of individuals who stutter, as seen in this study and previous studies (e.g., Arnold et al., 2016; Hughes et al., 2010; Hughes et al., 2017; Young & Byrd, 2024) and Byrd et al. (2024a; primary analysis, supplemental replication study). Across studies, untrained observers who are personally familiar with an adult who stutters rate more favorably than observers who reported they did not know a person who stutters. This finding is consistent with the well-documented contact theory wherein individuals with first-hand knowledge of a person from a specific identity or culture view these individuals more favorably (e.g., Allport, 1958; Pettigrew & Tropp, 2006). Findings suggest that rater familiarity should continue to be assessed, and controlled, when examining perceptions of the general public towards people who stutter.

## Limitations and Future Studies

The present study is not without limitations. First, although findings were replicated in two different settings, communication samples were collected in virtual space via Zoom.

Although virtual interviews and presentations are increasingly commonplace, it is possible that ratings of communication competence would be higher (or lower) if these interactions were observed in-person. Second, it was necessary to limit the number of videos included in the study due to the increased number of contexts and participants. As such, untrained observers rated only one post-treatment sample collected approximately one week following the final session.

Additional follow-up ratings are necessary to examine the long-term effects of treatment, as well as the inclusion of control conditions (e.g., non-stuttering adults, waitlist control) was included in the present assessment. Finally, we acknowledge that a variety of additional factors influence rater evaluation of any speaker (e.g., enthusiasm about topic, attire, perceived attractiveness, demographics of participant versus demographics of rater). The present study was large enough to let these factors freely vary across participants and across untrained observers (*N* = 106 to 117 per participant), and minimize the likelihood that these factors drive the outcomes in a systematic manner. Nevertheless, additional participant- and listener-based factors not measured in the present study cannot be ruled out as influential and should remain an area of examination in future studies.

## Conclusion

The purpose of this study was to replicate and extend a previous social validation study examining the treatment outcomes of the CARE Model. Findings replicated previous findings that positive gains in communication competence were observed by untrained observers in the absence of changes in stuttering severity. Findings also suggest that these effects were not limited to dyadic interactions but extend to oral presentations. Combined, outcomes of this large-scale study serve as social validation of a non-ableist approach designed to improve communication while explicitly excluding fluency goals and, instead, emphasizing stuttering openly as a fundamental step in effective communication.

## Data Availability

All data produced in the present study are available upon reasonable request to the authors

## Acknowledgements

This project was supported by the foundational grant support funded to the Arthur M. Blank Center for Stuttering Education and Research and endowed support provided through the Michael and Tami Lang Stuttering Institute, the Dr. Jennifer and Emanuel Bodner Developmental Stuttering Laboratory, and the Dealey Family Foundation Stuttering Clinic awarded to the second author. The authors thank Michael Mahometa for his assistance with statistical analyses. We would also like to thank the adult participants who stutter, as well as their families, who continue to participate in our ongoing clinical research.

## Appendix A

### Description of Treatment

Communication competence training serves as one of the four distinct components of the Blank Center CARE^™^ Model (Communication, Advocacy, Resilience, and Education). A brief summary of the 11-week (i.e., 22-session) manualized treatment protocol is provided below.

Treatment consists of two 60-minute sessions per week consisting of one group session as well as one individual session. Training provided during the individual sessions provide an opportunity to review what will be covered, prepare for the activities, and debrief for the weekly group sessions, wherein participants work towards strengthening their communication skills across distinct speaking scenarios, including mock job interviews, small group interactions, impromptu icebreakers, one-on-one interactions with unfamiliar persons, and multiple presentations varied both in purpose (e.g., informative, persuasive, inspirational) and audience composition (e.g., small and large groups, familiar and unfamiliar listeners).

- **Week 1** (Sessions 1 and 2): Participants are introduced to the competencies that comprise effective communication (i.e., language use, language organization, speech rate, intonation, volume, gestures, body position, eye contact, facial affect). Participants receive focused training in effective **body positioning and gestures.** Participants complete self-ratings of these communication competencies following an impromptu small group presentation.
- **Week 2** (Sessions 3 and 4): Participants identify and describe core components of communication competence and receive focused training in effective use of **facial affect,** while simultaneously continuing to strengthen competencies addressed in the prior sessions. Participants complete self-ratings of communication competencies following a small group informative speech presentation to unfamiliar persons.
- **Week 3** (Sessions 5 and 6): Participants identify and describe core components of communication competence and receive focused training in **turn-taking and listener awareness,** while simultaneously continuing to strengthen competencies addressed in prior sessions. Participants complete self-ratings of their communication competencies following impromptu dyadic exchanges as well as impromptu small group presentations to unfamiliar persons.
- **Week 4** (Sessions 7 and 8): Participants identify and describe core components of communication competence and receive focused training in effective **vocal variety (i.e., volume, rate, intonation),** while simultaneously continuing to strengthen competencies addressed in prior sessions. Participants complete self-ratings of communication competencies following impromptu dyadic exchanges, as well as impromptu small group presentations to unfamiliar persons.
- **Week 5** (Sessions 9 and 10): Participants identify and describe core components of communication competence and receive focused training in effective **language use and organization**, while simultaneously continuing to strengthen competencies addressed in prior sessions. Participants complete self-ratings of communication competencies after completing 10+ dyadic interactions with unfamiliar persons.
- **Week 6** (Sessions 11 and 12): Participants identify and describe core components of communication competence and receive additional focused training in **stuttering openly**- that is, making no attempts to avoid stuttering, increase fluency, and/or modify moments of stuttering. Participants complete self-ratings of communication competencies following an open mic presentation in a public forum.
- **Week 7** (Sessions 13 and 14): Participants **review and practice all core components of communication competence simultaneously**. Participants complete self-ratings of communication competencies following a persuasive speech given to a small group and a large group of unfamiliar persons.
- **Week 8** (Sessions 15 and 16): Participants **review and practice all core components of communication competence**. Participants complete self-ratings of communication competencies after serving as an interviewee in a series of panel interviews, with potential employers across diverse professions.
- **Week 9** (Sessions 17 and 18): Participants **review and practice all core components of communication competence.** Participants complete self-ratings of communication competencies following impromptu dyadic exchanges as well as impromptu presentation to a small group of unfamiliar persons.
- **Week 10** (Sessions 19 and 20): Participants **review and practice all core components of communication competence**. Participants complete self-ratings of communication competencies following impromptu dyadic exchanges as well as impromptu presentation to a small group of unfamiliar persons.
- **Week 11** (Sessions 21 and 22): Participants **review and practice all core components of communication competence.** Participants complete a self-rating of communication competencies after providing a presentation to an audience of >200 people.

## Appendix B

*Description of Non-overlapping, Untrained Observers Who Rated Each Video Sample* (*Condition* x *Timepoint* x *Context*)

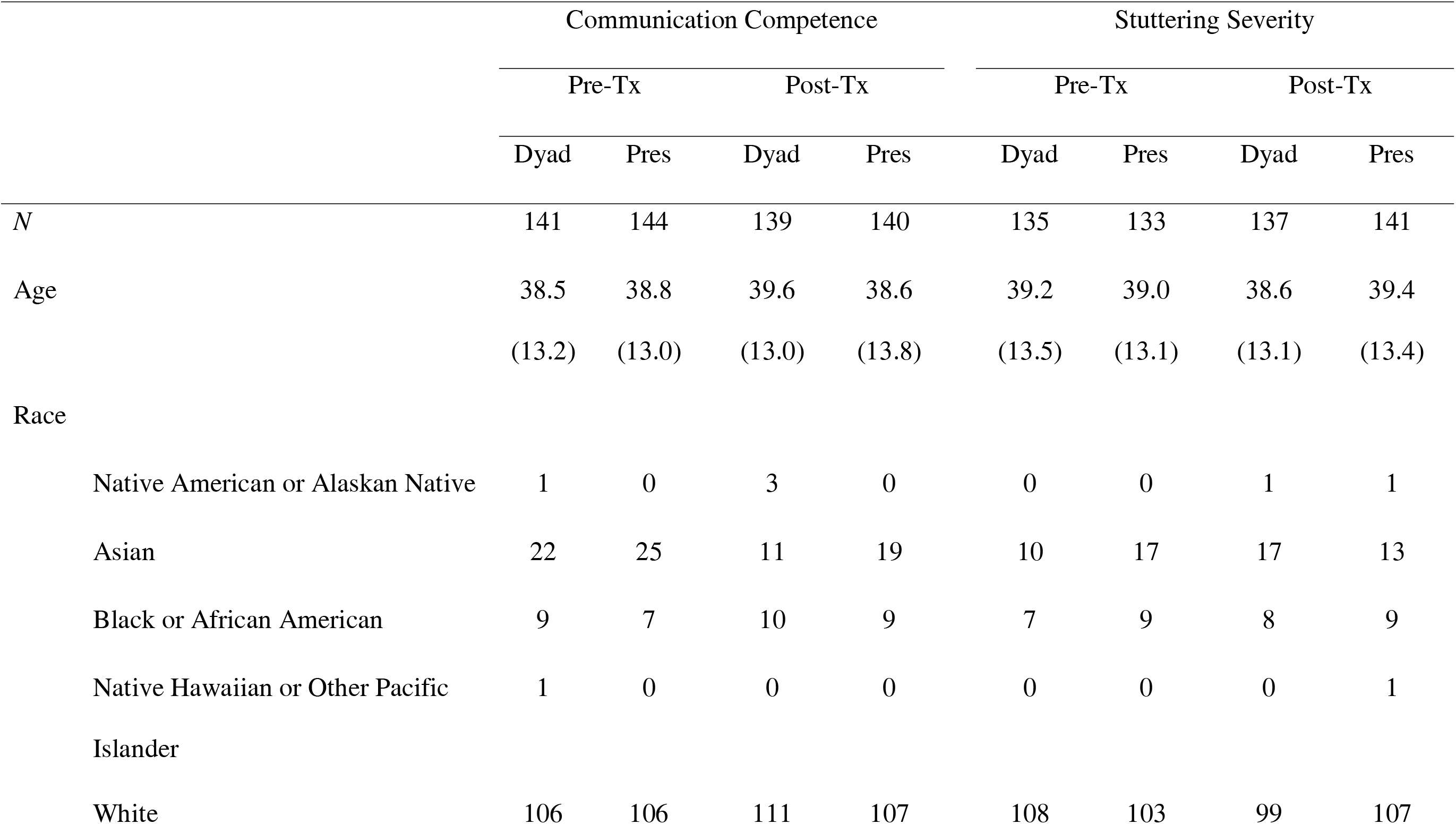
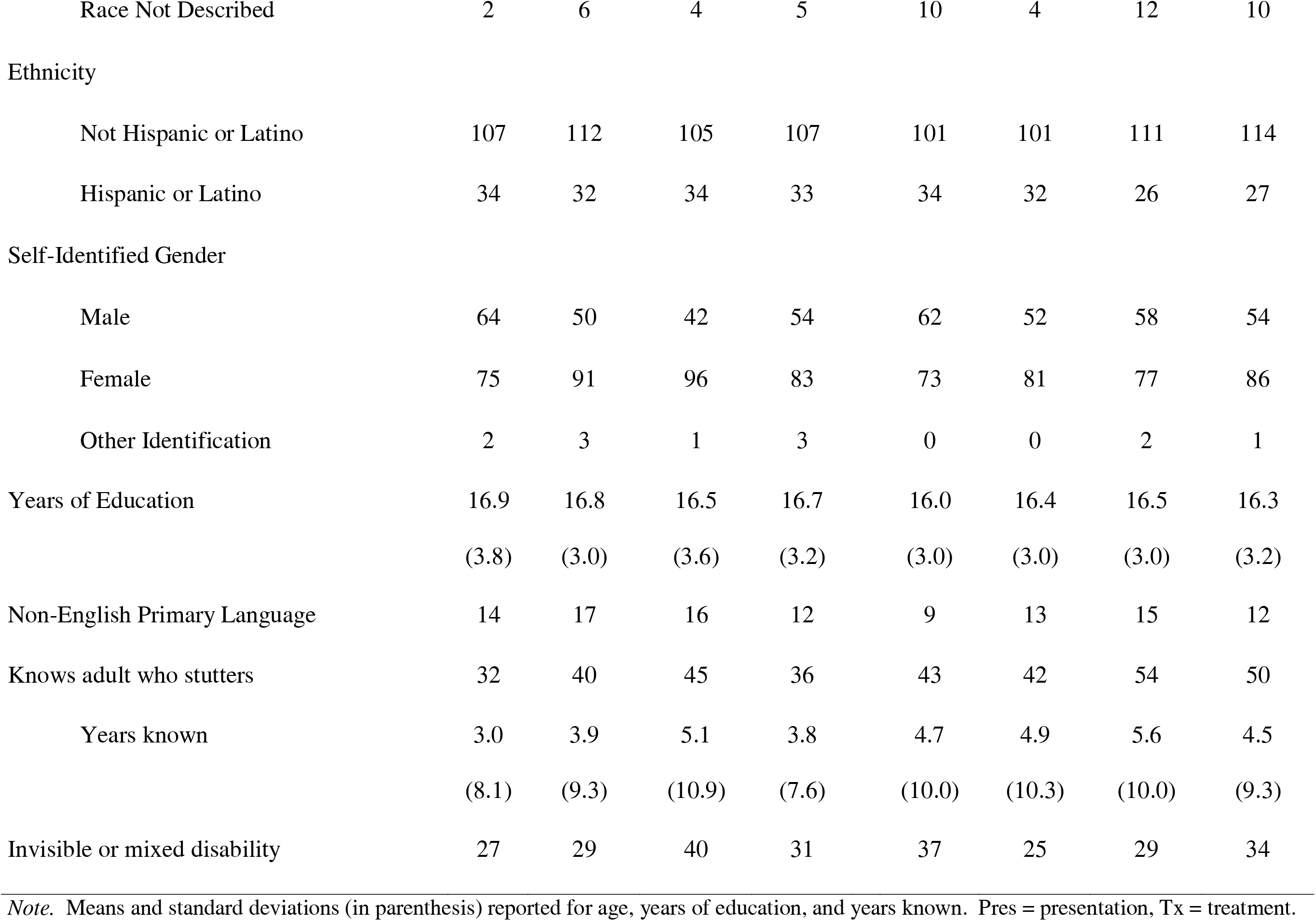

